# Sex-specific effects of birth weight on longitudinal behavioural outcomes in children and adolescents

**DOI:** 10.1101/2023.10.19.23297284

**Authors:** Lars Meinertz Byg, Carol Wang, John Attia, Craig Pennell

## Abstract

**Introduction:** Previous cross-sectional studies suggest that birth weight (BW) affects the development of aggression-, social- and attention problems differently in boys and girls. We sought to test if these differences could be confirmed in a longitudinal study.

**Methods:** The Raine Study provided prospectively collected data on perinatal variables and repeated child behaviour checklist assessments from ages five to seventeen. We used linear mixed effects model to determine crude and adjusted relationships between BW and childhood behaviour at a conservative significance threshold. Sensitivity analyses included an age ten teacher assessment.

**Results:** Data on behaviour, BW and sex, was available in 2269 participants. Male sex was associated with increased aggression problems at lower BW compared to females in the crude model (*β*: -0.436, 98.3%CI: [-0.844, - 0.0253]), but not the adjusted model (*β*: -0.310, 98.3%CI: [-0.742, 0.140]). Male sex was associated with increased attention problems at lower BW compared to females in both the crude model (*β*: -0.334, 98.3%CI: [- 0.530, -0.137]) and the adjusted model (*β*: -0.274, 98.3%CI: [-0.507, -0.0432]). Male sex was associated with increased social problems at lower BW compared to females in both the crude model (*β*: -0.164, 98.3%CI: [- 0.283, -0.0441]) and the adjusted model (*β*: -0.148, 98.3%CI: [-0.285, -0.00734]).

**Conclusion:** Using repeated measures from ages 5-17 we were able to show a crude and adjusted male vulnerability to lower BW in the development of attention problems and social problems. We did not find a BW x sex interaction for the development of aggressive behaviour.

## Introduction

Pre- and perinatal events can affect neurodevelopment with consequences for childhood and adult behaviour and mental illness[1]. Low birth weight (LBW) is a well-studied risk factor in the field of developmental origin of health and disease (DOHaD)[2] and predicts adult psychopathology[3–5]. Similarly, childhood behaviour has been associated with LBW in multiple settings, including twin studies [6–9]. The effects of birth weight (BW) on offspring behaviour have been reported as being dependent on offspring sex in non-related population settings, but there are inconsistencies in the directionality of these effects[10–12].

Murray et al., who examined the childhood behaviour checklist (CBCL) attention problems syndrome[13, 14], found a predominantly female vulnerability to LBW. In this study, 3700 individuals from the Brazilian PELOTAS birth cohort were followed prospectively with confounders recorded during pregnancy and behavioural assessment at age four. In contrast, Momany et al., using the DSM-IV ADHD Rating Scale and the Conners’ Rating Scale–Revised Short Form, found a male vulnerability for externalising and ADHD behaviour at age 12[12] in 900 individuals from North America. In this study, parents recalled confounders and BW, giving rise to the risk of recall bias. The most recent and high-powered study by Dooley et al. used an agnostic approach to examine the sex-specific effects of BW with the CBCL [14]. In this paper, nearly 10 000 individuals in the ABCD cohort were assessed at age 11 with subsequent testing of the association between BW and CBCL subscales- and total score[10]. At a conservative significance threshold, Dooley et al. found an increase in total CBCL scores, driven by sex differences in attention problems and aggression problems [10]; additionally, social problems had a nominally significant increase in males with LBW compared to females, but the sex interaction was insignificant after correction for multiple testing. A limitation of the ABCD study is the reliance on parental recollection of BW and potential confounders and the single behaviour assessment at age 11. The cross-sectional nature of previous studies limits conclusions regarding sex differences as low BW and biological sex are also implicated as determinants of behavioural phenotype trajectories across childhood and adolescence [15, 16]. The conflicting results and limitations cited above leave the question of a sex difference unresolved.

This paper aims to test sex differences in the relationship between BW and outcomes of aggression-, social- and attention problems. We seek to add to previous knowledge by using repeated measures across childhood and adolescence and to adjust for confounders collected during pregnancy to avoid recall bias.

## Participants and methods

### Study population

We analysed data from the Raine Study (https://rainestudy.org.au/) [17]. The Raine Study is a longitudinal study following mother-baby dyads recruited at or around 18 weeks gestation (n=2979) through the public antenatal clinic at King Edward Memorial Hospital and nearby private clinics in Perth, Western Australia, from May 1989 to November 1991. Offspring were followed up throughout childhood with modest attrition of mothers with lower age, education, income and non-European ancestry [18, 19]. The Human Research Ethics Committees at the University of Western Australia, King Edward Memorial Hospital, and Princess Margaret Hospital in Perth, Australia, granted ethics approval for each follow-up in the study.

### Outcome variables

Using the Achenbach System of Empirical Assessment (ASEBA) CBCL for Ages 4–18 (CBCL/4–18), we derived scores for attention problems, aggressive behaviour and social problems at ages five, eight, ten, 14 and 17. The CBCL/4–18 is a commonly used dimensional measure of child behaviour during the previous six months. The complete questionnaire contains 118 items and shows good internal reliability and validity in several population settings [13]. Participants were excluded from the analysis if they were missing more than eight items on the entire CBCL [20]. The attention problem subscale measures both problems of attention, impulsivity and hyperactivity and consists of 11 items (score 0-22). The social problems scale measures peer interaction problems and consists of 8 items (score 0-16). The aggressive behaviour scale consists of 20 items (score 0-40). The CBCL is a highly validated psychometric tool in many countries and is used in the clinical setting as a guide and screening tool[13, 14, 21]. Previous authors have used the CBCL raw scores (and not T-score corrected for age and sex) to examine sex differences[10, 11]. We chose the same approach of regressing the raw scores. The CBCL syndrome scores across populations tend to be right skewed and the clinical cut-offs for the raw scores are therefore low (for attention problems clinical relevance is assumed around 7 to 8 points depending on sex and age). The primary aim of our paper was the sex x BW interaction (to answer the question of a differential effect between males and females), and to account for the reductions in CBCL raw scores seen with increasing age, we added age at assessment as a fixed term to our regression (see below).

We used two additional instruments for sensitivity analyses: the 1991 ASEBA preschool form of the CBCL (also filled out by parents) and the Teacher Report Form (TRF). The preschool CBCL scale for aggressive behaviour was assessed at two years of age and had 33 items (score 0-66). The 1991 edition of the preschool questionnaire did not have a validated attention problem and social problem scale. The TRF is another well-validated form under ASEBA and can be used in the clinical setting to support findings from the CBCL[22]. The TRF attention problems scale has 20 items (score 0-40), the aggression problems scale has 25 items (score 0-50) and the social problems scale has 13 items (score 0-26)

### Early life determinants and potential confounders

Gestational age (GA) was determined either by the date of the last menstrual period (LMP) or fetal biometry at the 18-week gestation ultrasound (USS) examination. BW and fetal sex were retrieved from hospital records. Different populations have different normal spectra for BW[23], and we used the normalised BW as the exposure.

Information on confounders was recorded at prenatal visits at gestational week 18 by maternal questionnaire. We chose only to include factors that might be associated with both BW and behavioural outcomes, meaning no post-natal variables were used. We did, however, include the cohort age at assessment as a fixed term to minimise the noise from age-related CBCL-score reductions. Potential confounders were inserted in a directed acyclic graph (supplementary Figure 1) to help us decide on the co-variables to include in our models. Smoking, alcohol consumption, economic class and maternal education were recorded as ordinal 5-6 level variables and were treated as continuous in the multivariable analysis.

### Statistics

All statistical analyses and graphs were performed in “R” [24] and its associated libraries “Gmisc”, “lme4”, “lmeresampler”, “lmertest” and “boot”. The Wilcoxon rank-sum test was used to compare sample distributions in the continuous baseline variables and CBCL outcomes between males and females.

Testing the associations between BW and behavioural outcomes was done with mixed-effects modelling to avoid pseudo-replication from repeated measurements of the same participant. Recent work has demonstrated that the treatment of ordinal data as continuous does not impact inference in most situations [25], and linear mixed modelling is robust to missing data and violation of distributional assumptions[26]. We, therefore, chose to treat the CBCL scores as continuous variables. As we examined three different (albeit correlated) outcomes, we applied a Bonferroni-correction of 3 for our alpha, meaning statistical significance was set at 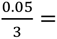 0.01667.

Model diagnostics were evaluated by examining histograms and qq-plots of residuals and random effects. CBCL subscale scores are highly right-skewed, and our residuals displayed significant violations of distributional assumption. We, therefore, performed a non-parametric bootstrap at the participant-ID level with 5000 simulations, as suggested by Thai et al. [27], to derive estimates and 98.3 % confidence intervals, which were then used for primary inference. Approximate p-values were calculated from the bootstrapped estimate z-statistic. For the sensitivity analysis using the TRF (see below) we performed a simple linear regression of TRF-scores as described below, and subsequently did a non-parametric bootstrap with 5000 simulations because of non-normality in our residuals.

We had 4 model levels, with the sex - interaction (*B_3_*) being the variable of primary interest. If *y_IA_* is the CBCL score for a given individual at a given assessment age, *ε_IA_* is the error term, *B*_0_ is the intercept and *μ_I_*_0_ is the random effect on the intercept, our models were as follows:

1. An unadjusted model including only a fixed effect of BW (*B*_1_).
2. An unadjusted model as in 1) but with a fixed effect of sex (*B*_2_) and a sex interaction term (*B*_3_\**sex_IA_*) where females were the reference category,
3. A model as in 2), but with DAG-determined confounders (*B_conv_*) and the age at assessment (*B*_4_) added as fixed effects
4. A parsimonious DAG-determined model with main effects and interaction. The parsimonious model was derived using backward variable removal from the full model 3) by examining p-values from the linear mixed effects model. Only variables with a p-value < 0.2 were included in model 4)

The full model, including interactions and confounders (model 3 and 4) was specified as follows:

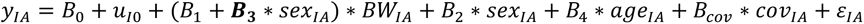

For prespecified sensitivity analyses, we excluded preterm births (less than 37 weeks gestational age at birth); additionally, aggressive behaviour was reassessed with the inclusion of an age two behavioural assessment using the preschool form of the CBCL. We added a fixed term to the model to account for the increased number of items on the preschool aggression problems. Finally, as parent characteristics have an association with both BW and CBCL scores, it is possible that parents of children with lower BW could rate behaviour differently and bias the estimate of childhood phenotype; therefore, we wanted to ensure that there was no disagreement between teacher ratings at age ten and primary findings from longitudinal parent ratings.

We used the STROBE cohort checklist when writing our paper[28].

## Results

In the Raine Study, 2868 (100 %) live births were assessed for eligibility (Supplementary Figure 2). If a mother had more than one offspring participate in the Raine Study (multiple gestation or siblings from distinct pregnancies), the younger offspring were excluded for a total of 2731 participants (95.2%). We then identified the 2276 participants with at least one valid CBCL assessment from ages 5-17 (79.4 %) (Table 1). BW and biological sex were recorded for 2269 (79.1 %) participants (used in the model 1 + model 2 regression). Information on DAG-determined confounders was available for 1994 (69.5 %) participants (used in the model 3 regression). The backward variable removal for the attention- and social problem scale did not yield any additional participants for analysis, but gestational age was dropped from the aggressive behaviour parsimonious model 4 for a total of 1996 (69.6 %) participants. Maternal baseline variables were evenly distributed between pregnancies with male and female offspring (Table 1). The mean BW in the Raine Study cohort was 3318.5 g (1-SD = 594.5 g) and males had higher BW than females. Males had higher CBCL scores across the examined subscales including teacher assessments and preschool assessments (Table 2 and supplementary Table 3). CBCL follow-up decreased as the cohort age increased, with substantial attrition at age 17. Compared to dropouts, the Raine Study participants with at least 1 CBCL assessment had more favourable socio-economic status, less pregnancy-related risk behaviour and a higher BW (supplementary Table 1).

**Table 1.** Baseline demographics for the analytic cohort (n = 2269)

|  | Female (n = 1095) | Male (n = 1174) |
| --- | --- | --- |
| **Birth weight (g)** |  |  |
|  | 3251.1 (±586.0) | 3381.4 (±595.6) |
| **Maternal age at birth (years)** |  |  |
| Mean (SD) | 28.6 (±5.9) | 28.5 (±5.7) |
| Missing | 0 (0%) | 1 (0.1%) |
| **Income level (AUD)†** |  |  |
| Mean (SD) | 3.7 (±1.2) | 3.7 (±1.2) |
| Missing | 68 (6.2%) | 50 (4.3%) |
| **Maternal body mass index (kg/m<sup>2</sup>)** |  |  |
| Mean (SD) | 22.3 (±4.3) | 22.4 (±4.2) |
| Missing | 0 (0%) | 2 (0.2%) |
| **Maternal race** |  |  |
| European descent | 972 (88.8%) | 1,058 (90.1%) |
| Aboriginal | 15 (1.4%) | 16 (1.4%) |
| Polynesian | 10 (0.9%) | 8 (0.7%) |
| Vietnamese | 2 (0.2%) | 4 (0.3%) |
| Chinese | 50 (4.6%) | 49 (4.2%) |
| Indian | 32 (2.9%) | 27 (2.3%) |
| Other | 14 (1.3%) | 11 (0.9%) |
| Missing | 0 (0.0%) | 1 (0.1%) |
| **Maternal education††** |  |  |
| Mean (SD) | 1.2 (±1.5) | 1.2 (±1.5) |
| Missing | 0 (0%) | 1 (0.1%) |
| **Diabetes or hypertension in pregnancy** |  |  |
| Absent | 914 (83.5%) | 967 (82.4%) |
| Present | 181 (16.5%) | 206 (17.5%) |
| Missing | 0 (0.0%) | 1 (0.1%) |
| **Gestational age at birth (weeks)** |  |  |
| Mean (SD) | 38.8 (±2.2) | 38.8 (±2.1) |
| Missing | 1 (0.1%) | 1 (0.1%) |
| **Smoking in pregnancy†††** |  |  |
| Mean (SD) | 0.7 (±1.3) | 0.5 (±1.2) |
| Missing | 87 (7.9%) | 80 (6.8%) |
| **Treatment for psychiatric disorder** |  |  |
| Absent | 1,073 (98.0%) | 1,147 (97.7%) |
| Present | 22 (2.0%) | 26 (2.2%) |
| Missing | 0 (0.0%) | 1 (0.1%) |
| **Maternal alcohol consumption††††** |  |  |
| Mean (SD) | 4.8 (±1.4) | 4.8 (±1.3) |
| Missing | 0 (0%) | 1 (0.1%) |

†Family income: 1=Less than \$7,000, 2=\$7,000-\$11,999, 3=\$12,000-\$23,999, 4=\$24,000-\$35,000, 5=\$36,000 or more
††Education: 0 = None or `Other`, 1 = Trade certificate or apprenticeship, 2=Professional registration (non-degree), 3=College diploma or degree, 4=University degree
††† 0=None, 1=1 to 5 daily, 2=6 to 10 daily, 3=11 to 15 daily, 4=16 to 20 daily, 5=21 or more per day
††††1=Daily, 2=Several times per week, 3=Approximately once per week, 4, Less than once per week, 5=One binge effort, 6=Never

**Table 2.** Child behaviour checklist (CBCL) scores for the analytic cohort (n = 2269)

|  | Female (n = 1095) | Male (n = 1174) | P-value |
| --- | --- | --- | --- |
| **CBCL Attention problems age 5** |  |  | **< 0.0001** |
| Mean (SD) | 2.6 (±2.8) | 3.6 (±3.1) | |
| Missing | 97 (8.9%) | 114 (9.7%) |  |
| **CBCL Attention problems age 8** |  |  | **< 0.0001** |
| Mean (SD) | 2.5 (±2.9) | 3.7 (±3.5) | |
| Missing | 134 (12.2%) | 157 (13.4%) |  |
| **CBCL Attention problems age 10** |  |  | **< 0.0001** |
| Mean (SD) | 2.0 (±2.8) | 3.2 (±3.4) | |
| Missing | 173 (15.8%) | 181 (15.4%) |  |
| **CBCL Attention problems age 14** |  |  | **< 0.0001** |
| Mean (SD) | 1.9 (±2.6) | 2.8 (±3.2) | |
| Missing | 265 (24.2%) | 307 (26.1%) |  |
| **CBCL Attention problems age 17** |  |  | **0.0007** |
| Mean (SD) | 1.6 (±2.4) | 2.1 (±2.7) | |
| Missing | 443 (40.5%) | 512 (43.6%) |  |
| **CBCL Aggression problems age 5** |  |  | **< 0.0001** |
| Mean (SD) | 7.8 (±5.7) | 9.5 (±6.7) | |
| Missing | 97 (8.9%) | 114 (9.7%) |  |
| **CBCL Aggression problems age 8** |  |  | **< 0.0001** |
| Mean (SD) | 6.6 (±5.7) | 8.5 (±6.9) | |
| Missing | 134 (12.2%) | 157 (13.4%) |  |
| **CBCL Aggression problems age 10** |  |  | **< 0.0001** |
| Mean (SD) | 5.4 (±5.2) | 7.1 (±6.3) | |
| Missing | 173 (15.8%) | 181 (15.4%) |  |
| **CBCL Aggression problems age 14** |  |  | **0.036** |
| Mean (SD) | 5.2 (±5.6) | 5.9 (±6.1) | |
| Missing | 265 (24.2%) | 307 (26.1%) |  |
| **CBCL Aggression problems age 17** |  |  | **0.75** |
| Mean (SD) | 3.8 (±4.9) | 3.9 (±4.9) | |
| Missing | 443 (40.5%) | 512 (43.6%) |  |
| **CBCL Social problems age 5** |  |  | **0.033** |
| Mean (SD) | 1.6 (±1.7) | 1.8 (±1.9) | |
| Missing | 97 (8.9%) | 114 (9.7%) |  |
| **CBCL Social problems age 8** |  |  | **0.11** |
| Mean (SD) | 1.6 (±1.8) | 1.8 (±2.2) | |
| Missing | 134 (12.2%) | 157 (13.4%) |  |
| **CBCL Social problems age 10** |  |  | **0.0003** |
| Mean (SD) | 1.4 (±2.0) | 1.8 (±2.2) | |
| Missing | 173 (15.8%) | 181 (15.4%) |  |
| **CBCL Social problems age 14** |  |  | **0.077** |
| Mean (SD) | 1.1 (±1.8) | 1.3 (±2.0) | |
| Missing | 265 (24.2%) | 307 (26.1%) |  |
| **CBCL Social problems age 17** |  |  | **0.93** |
| Mean (SD) | 0.8 (±1.5) | 0.7 (±1.4) | |
| Missing | 443 (40.5%) | 512 (43.6%) |  |

P-value calculated by Willcoxon rank-sum test

There was no significant main effect of BW on aggressive behaviour in the unadjusted model 1(*β*: -0.0872, 98.3%CI: [-0.294, 0.130]) (Table 3). We found a significant sex x BW interaction from male sex in the unadjusted model 2 (*β*: -0.436, 98.3%CI: [-0.844, -0.0253]). The sex x BW interaction diminished after confounder adjustment in our complete (*β*: -0.315, 98.3%CI: [-0.744, 0.127]) model 3 and parsimonious (*β*: - 0.310, CI: [-0.742, 0.140]) model 4 and did not show any significant interaction between BW and male sex. (Table 3)

**Table 3.** The association between BW and aggression problems in the Raine Study ages 5-17.

|  | Univariable model<br>(Model 1) | Crude sex-interaction<br>(Model 2) | Fully adjusted<br>(Model 3)** | Parsimonious model<br>(Model 4)*** |
| --- | --- | --- | --- | --- |
| **Main effect** | $\beta$: -0.0872<br>CI*: [-0.294, 0.130]<br>SE: 0.089<br>P-value*: 0.33 | NA | NA | NA |
| **Baseline effect**<br>**(females)** | NA | $\beta$: 0.0602<br>CI*: [-0.191, 0.315]<br>SE: 0.106<br>P-value*: 0.57 | $\beta$: -0.00802<br>CI*: [-0.386, 0.380]<br>SE: 0.161<br>P-value*: 0.96 | $\beta$: -0.0160<br>CI*: [-0.335, 0.298 ]<br>SE: 0.133<br>P-value*: 0.90 |
| **Sex Interaction**<br>**(males)** | NA | $\beta$: **-0.436**<br>CI*: **[-0.844, -0.0253]**<br>SE: **0.172**<br>P-value*: **0.011** | $\beta$: -0.315<br>CI*: [-0.744, 0.127]<br>SE: 0.182<br>P-value*: 0.08 | $\beta$: -0.310<br>CI*: [-0.742, 0.140]<br>SE: 0.185<br>P-value*: 0.09 |

\* P-value is approximated based on Z-statistic of the bootstrapped SE, but significance is derived from the 98.3 % CI
\*\* Adjusted for age at assessment, maternal BMI, maternal education,maternal psychiatric illness, gestational age at birth, maternal age at birth, maternal smoking during pregnancy, Diabetes mellitus or hypertension in pregnancy, family income during pregnancy, maternal ethnicity and maternal alcohol consumption during pregnancy.
\*\*\* Adjusted for age at assessment, maternal BMI, maternal age at birth, maternal smoking during pregnancy, maternal alcohol consumption, family income level and maternal race

There was a significant inverse linear effect of BW on attention problems in the unadjusted model 1 (*β*: -0.131, 98.3%CI: [-0.227, -0.0252]) with a 1 SD increase in BW reducing attention problems by 0.139 points (Table 4). We found a significant sex x BW interaction in the unadjusted model 2 (*β*: -0.334 98.3%CI: [-0.530, -0.137]) with male sex driving the association between BW and attention problems in the overall population. The sex interaction was robust to multivariable adjustment in the complete model 3 (*β*: -0.276 98.3%CI: [-0.503, - 0.0388]) and in the parsimonious model 4 (*β*: -0.274 98.3%CI: [-0.507, -0.0432]) (Table 4)

**Table 4.**
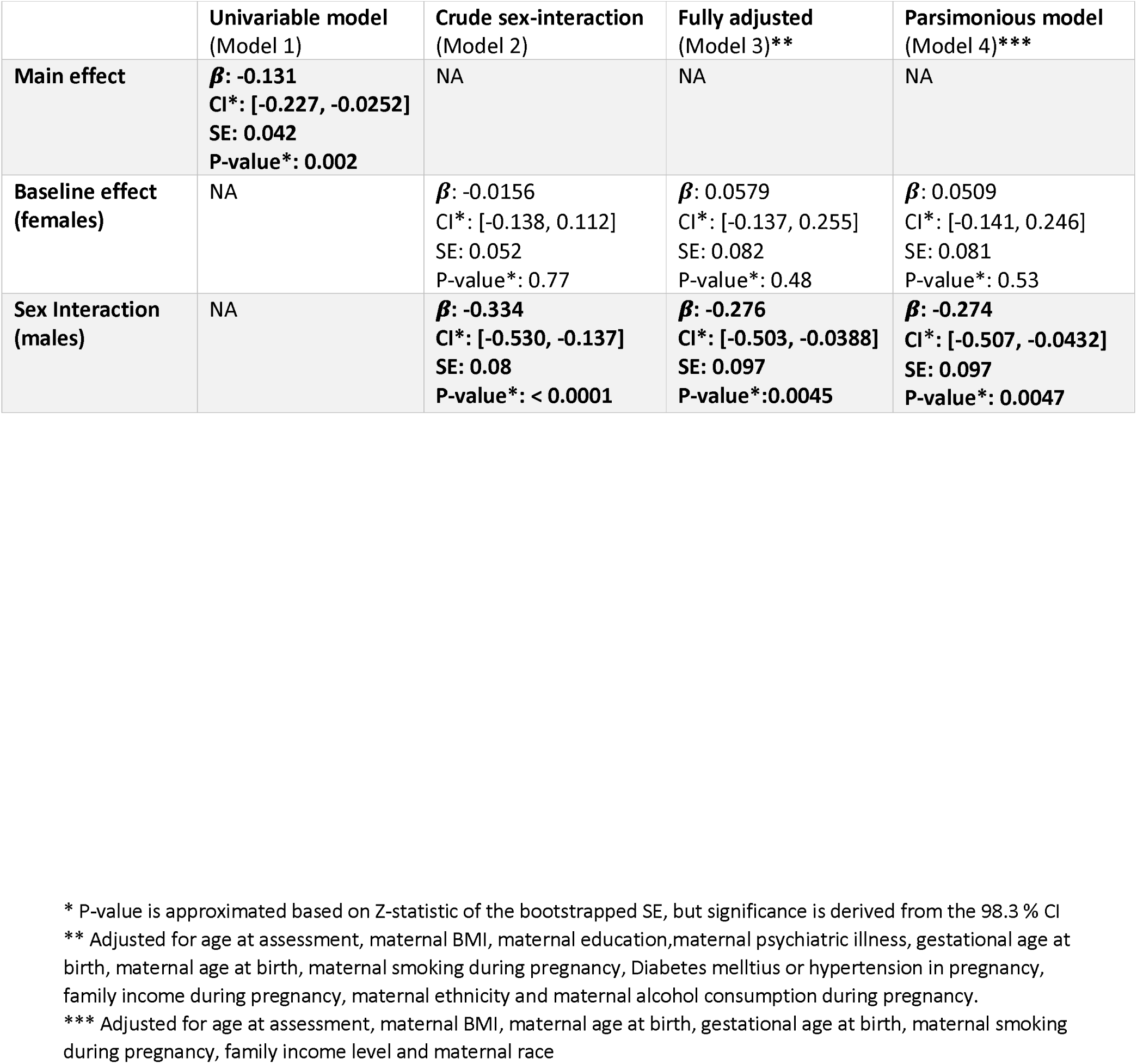
The association between BW and attention problems in the Raine Study ages 5-17.

|  | Univariable model<br>(Model 1) | Crude sex-interaction<br>(Model 2) | Fully adjusted<br>(Model 3)** | Parsimonious model<br>(Model 4)*** |
| --- | --- | --- | --- | --- |
| **Main effect** | **$\beta$: -0.131**<br>**CI\*: [-0.227, -0.0252]**<br>**SE: 0.042**<br>**P-value\*: 0.002** | NA | NA | NA |
| **Baseline effect**<br>**(females)** | NA | **$\beta$: -0.0156**<br>**CI\*: [-0.138, 0.112]**<br>**SE: 0.052**<br>**P-value\*: 0.77** | **$\beta$: 0.0579**<br>**CI\*: [-0.137, 0.255]**<br>**SE: 0.082**<br>**P-value\*: 0.48** | **$\beta$: 0.0509**<br>**CI\*: [-0.141, 0.246]**<br>**SE: 0.081**<br>**P-value\*: 0.53** |
| **Sex Interaction**<br>**(males)** | NA | **$\beta$: -0.334**<br>**CI\*: [-0.530, -0.137]**<br>**SE: 0.08**<br>**P-value\*: < 0.0001** | **$\beta$: -0.276**<br>**CI\*: [-0.503, -0.0388]**<br>**SE: 0.097**<br>**P-value\*: 0.0045** | **$\beta$: -0.274**<br>**CI\*: [-0.507, -0.0432]**<br>**SE: 0.097**<br>**P-value\*: 0.0047** |

\* P-value is approximated based on Z-statistic of the bootstrapped SE, but significance is derived from the 98.3 % CI
\*\* Adjusted for age at assessment, maternal BMI, maternal education, maternal psychiatric illness, gestational age at birth, maternal age at birth, maternal smoking during pregnancy, Diabetes mellitus or hypertension in pregnancy, family income during pregnancy, maternal ethnicity and maternal alcohol consumption during pregnancy.
\*\*\* Adjusted for age at assessment, maternal BMI, maternal age at birth, gestational age at birth, maternal smoking during pregnancy, family income level and maternal race

There was a significant inverse linear effect of BW on social problems in the unadjusted model 1(*β*: -0.0646 98.3%CI: [-0.123 -0.0043]) with a 1 SD increase in BW reducing social problems by 0.0646 points (Table 5). We found a significant sex x BW interaction from male sex in the unadjusted model 2 (*β*: -0.164 98.3%CI: [- 0.283, -0.0441]), with male sex driving the association between BW and social problems in the overall population. The sex interaction was robust to multivariable adjustment in the complete model 3 (*β*: -0.149 98.3%CI: [-0.286, -0.013]) and the parsimonious model 4 (*β*: -0.148 98.3%CI: [-0.285, -0.00734]) (Table 5)

**Table 5.** The association between BW and social problems in the Raine Study ages 5-17.

|  | Univariable model<br>(Model 1) | Crude sex-interaction<br>(Model 2) | Fully adjusted<br>(Model 3)** | Parsimonious model<br>(Model 4)*** |
| --- | --- | --- | --- | --- |
| Main effect | $\beta$: -0.0646<br>CI*: [-0.123 -0.0043]<br>SE: 0.025<br>P-value*: 0.0096 | NA | NA | NA |
| Baseline effect<br>(females) | NA | $\beta$: 0.00839<br>CI*: [-0.075, 0.093]<br>SE: 0.035<br>P-value*: 0.81 | $\beta$: 0.0663<br>CI*: [-0.0574, 0.193]<br>SE: 0.052<br>P-value*: 0.21 | $\beta$: 0.0617<br>CI*: [-0.0601, 0.184]<br>SE: 0.051<br>P-value*: 0.23 |
| Sex Interaction<br>(males) | NA | $\beta$: -0.164<br>CI*: [-0.283, -0.0441]<br>SE: 0.05<br>P-value*: 0.001 | $\beta$: -0.149<br>CI*: [-0.286, -0.013]<br>SE: 0.058<br>P-value*: 0.009 | $\beta$: -0.148<br>CI*: [-0.285, -0.00734]<br>SE: 0.058<br>P-value*: 0.011 |

\* P-value is approximated based on Z-statistic of the bootstrapped SE, but significance is derived from the 98.3 % CI
\*\* Adjusted for age at assessment, maternal BMI, maternal education, maternal psychiatric illness, gestational age at birth, maternal age at birth, maternal smoking during pregnancy, diabetes mellitus or hypertension in pregnancy, family income during pregnancy, maternal ethnicity and maternal alcohol consumption during pregnancy.
\*\*\* Adjusted for age at assessment, maternal BMI, maternal psychiatric illness, maternal age at birth, gestational age at birth, maternal smoking during pregnancy, maternal alcohol consumption during pregnancy and family income level

For sensitivity analyses of aggressive behaviour, we had CBCL data from the age two round of assessments for aggressive behaviour. Inclusion yielded an additional 115 participants for regression (model 1 n= 2384). A full reanalysis with inclusion of age 2 assessments did not change the estimate size or significance (supplementary Table 3).

Exclusion of preterm births (n = 147) had little effect on the estimate parameters for social- and attention problems (supplementary Table 4). We saw a 30 % reduction in the beta coefficient of the BW x sex interaction for aggressive behaviour.

Age ten teacher assessment of child behaviour with the TRF was available for 1585 individuals with relevant covariables for the model 4 regression. No sex x BW interaction reached significance at the 98.3 % level, but we saw directionally similar associations between BW and childhood assessments as was seen with the parent assessments (supplementary Table 5).

## Discussion

Using repeated parent ratings across childhood and adolescence we examined crude and confounder-adjusted sex differences in the association between BW and aggression, attention, and social problems from ages 5-17 years. We found longitudinal sex difference in the relationship between birth weight and attention problems and social problems, but not aggression problems.

We found no BW x sex interaction in aggressive behaviour and so could not reproduce the Dooley et al. findings showing a male vulnerability. Although there was a crude association in our model 2, the signal weakened with multiple regression in models 3 and 4. The inclusion of age two assessments, exclusion of preterm births or assessment by teachers did not change inference. Our findings also contrast with results from Momany et al. [12], who, similar to Dooley et al. found a BW x sex interaction. Both corrected for variables collected at the time of assessment. One explanation for this discrepancy could be that we did not have the power to detect a difference; however, our study population was more than double that of Momany et al. Alternatively, it is possible that the earlier findings were subject to residual confounding given that our models were comprehensively but not excessively adjusted with data collected during pregnancy and were consistent across the multiple models. Alcohol exposure and maternal BMI were not included in previous papers, and in the paper by Dooley et al. prenatal smoking was not included in multivariable modelling.

We found a BW x sex interaction in attention problems, confirming a male vulnerability. This finding was robust across all models. The magnitude of this effect was comparable to that seen in the Dooley paper (0.46 vs 0.35 points per kg BW). The exclusion of preterm births did not dramatically change the effect size. Sensitivity analysis using raw scores from the TRF was in directional agreement with CBCL scores. The TRF models were not significant at our Bonferroni corrected threshold; however, there was a signal when using the 95 % CI (data not shown). Although not conclusive, this data supports the results from the primary analysis. Our finding is also in line with the finding from Momany et al. [12]. Our finding is inconsistent with results from Murray et al., who found a female vulnerability in the Brazilian birth cohort PELOTAS [29]. It is not clear why our results show opposite findings, but there are differences between our designs that might contribute. First, Murray et al. used a very early behaviour assessment (age 4), and males and females have been known to manifest attention problems differently across childhood and adolescence[29]. Second, they used categorical exposures (low vs appropriate birth weight) with a 2500 g threshold for both males and females, although BW spectra differ between males and females. Third, they approached the CBCL as an ordinal outcome variable, whereas we treated it as a continuous outcome. Fourth, they had a markedly different study population compared to us, characterised by less affluent mothers with different pregnancy-related risk behaviours, which might change the relationship between BW and parent assessment. Our results also contrast with the apparent null finding from the sibling control study by Pettersson et al. [30]. Although not compared directly, they found a similar relationship between BW and rates of neurodevelopmental disorders diagnosed across life in males and females (supplementary Table 9 in [30]). It is important to note that “neurodevelopmental disorders” also encompass autism; furthermore, a diagnostic category is different from the continuous spectrum in the CBCL attention problem scale. Importantly, incidence of ADHD diagnosis peaks later in girls as compared to boys [31], which might suggest an age-dependent symptom manifestation. It is possible that the age 17 cut-off of in our data collection therefore represents a period of symptomatic latency for females.

We found a BW x sex interaction in social problems suggesting a male vulnerability. This finding was robust across all models. The exclusion of preterm births did not change the inference, and teacher assessment agreed with the results from the primary analysis. There have been previous reports of an overall association between lower birth weight and social problems in childhood and adulthood [32, 33]; however, Dooley et al were to our knowledge the first authors to examine a sex difference directly and did not find a significant sex difference. Our results suggest that lower birth weight increases social problems in males but given the novelty of this finding replication is needed.

This study’s strengths are the prospective data collection (avoiding recall bias), limited attrition, and use of multiple well-validated psychometric instruments. The models are also robust in using repeated measures on individuals and a careful approach to model construction using DAGs to make explicit the relationship between our variables. This study’s limitations are the primarily Caucasian population in the cohort and the age 17 cut-off for psychometric evaluation.

In conclusion, using repeated measures from ages 5-17 with correction of maternal baseline variables collected during pregnancy, we were able to show a male vulnerability of low birth weight in the development of attention problems and social problems; however, we failed to find a similar sex-interaction for the development of aggressive behaviour.

## Data Availability

All data used in the present study is available only through the Raine Study Steering comitee

## Acknowledgements and Funding Statement

We would like to acknowledge the Raine Study participants and their families for their ongoing participation in the study and the Raine Study team for study co-ordination and data collection. We also thank the NHMRC for their long term contribution to funding the study over the last 30 years. The core management of the Raine Study is funded by The University of Western Australia, Curtin University, Telethon Kids Institute, Women and Infants Research Foundation, Edith Cowan University, Murdoch University, The University of Notre Dame Australia and the Raine Medical Research Foundation. The Raine Study Gen2-14 year follow-up was sponsored by the NHMRC Program Grant 211912 and 003209. The Raine Study Gen2-17 year follow-up was sponsored by the NHMRC Program Grant 353514.

## Correspondence to

Dr. Lars Meinertz Byg, Lot 1 Kookaburra Cct, New Lambton Heights NSW 2305, 04 23215758

## Access to data and Data sharing

The first author of this paper had full access to all the data in the study and take responsibility for the integrity of the data and the accuracy of the data analysis.

## Conflict of interest

The authors declare no conflicts of interest.

## Notes

### Competing Interest Statement

The authors have declared no competing interest.

