## Supplemental materials for "Sex-specific effects of birth weight on longitudinal behavioural outcomes in children and adolescents"

### Supplementary Figure 1 – Directed Acyclic Graph

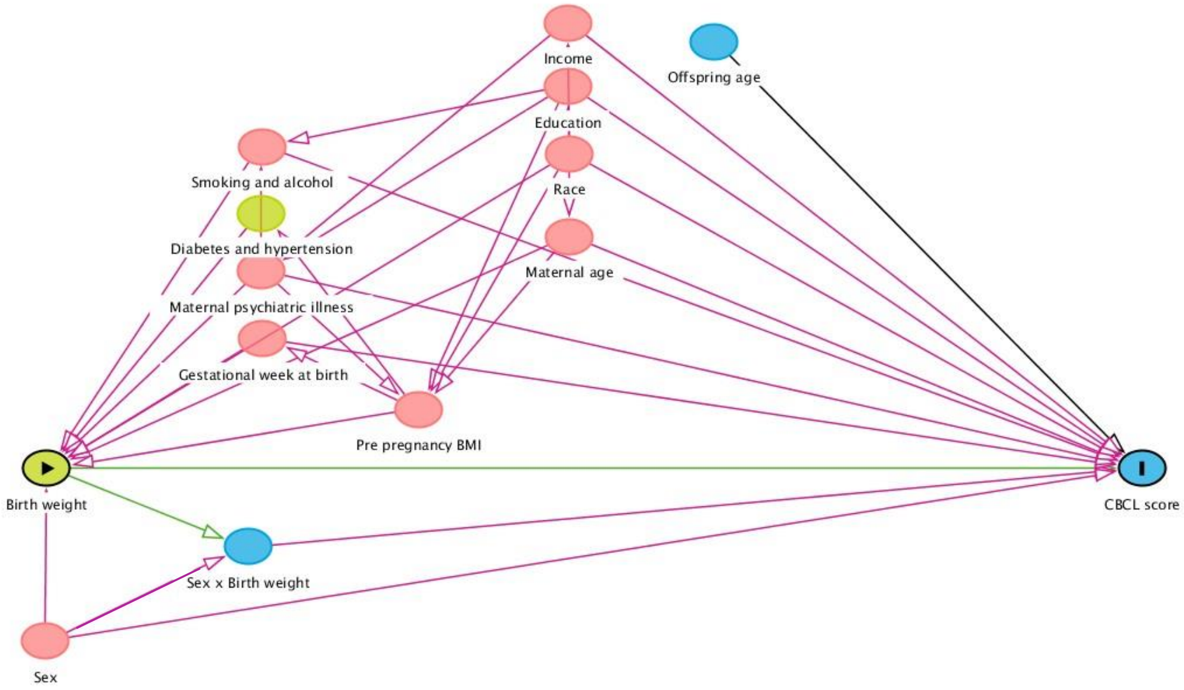


Figure 1. Conceptual framework for the primary analysis of sex-specific effects of birthweight on childhood behaviour with possible maternal and pregnancy-related confounders. **CBCL**: Child Behaviour Checklist, **BMI**: Body Mass Index.

### Supplementary Figure 2 – Flowchart of cohort selection

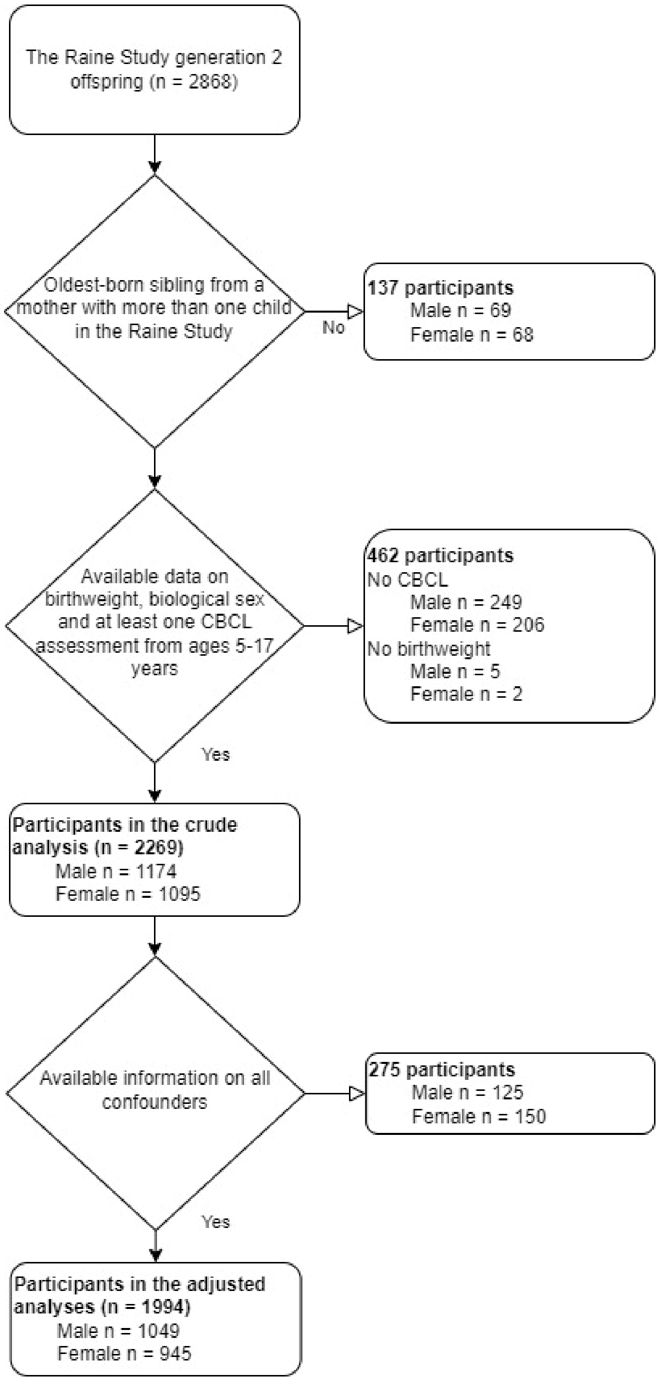


Figure 2. Flow diagram of cohort selection. **CBCL:** Child behaviour checklist 4/18 years

Supplementary Table 1. Demographics for analytic cohort vs excluded participants (n = 2868)

| | Excluded prticipants (n=599) | Analytic cohort (n=2269) |
|---|---|---|
| **Birth weight (g)** | | |
| Mean (SD) | 3159.0 (±702.2) | 3318.5 (±594.5) |
| Missing | 10 (1.7%) | 0 (0%) |
| **Maternal age at birth (years)** | | |
| Mean (SD) | 26.1 (±6.0) | 28.5 (±5.8) |
| Missing | 71 (11.9%) | 1 (0.0%) |
| **Income level (AUD)†** | | |
| Mean (SD) | 3.2 (±1.3) | 3.7 (±1.2) |
| Missing | 112 (18.7%) | 118 (5.2%) |
| **Maternal body mass index (kg/m^2)** | | |
| Mean (SD) | 22.4 (±4.6) | 22.3 (±4.2) |
| Missing | 63 (10.5%) | 2 (0.1%) |
| **Maternal race** | | |
| European descent | 442 (73.8%) | 2,030 (89.5%) |
| Aboriginal | 36 (6.0%) | 31 (1.4%) |
| Polynesian | 6 (1.0%) | 18 (0.8%) |
| Vietnamese | 2 (0.3%) | 6 (0.3%) |
| Chinese | 26 (4.3%) | 99 (4.4%) |
| Indian | 15 (2.5%) | 59 (2.6%) |
| Other | 9 (1.5%) | 25 (1.1%) |
| Missing | 63 (10.5%) | 1 (0.0%) |
| **Maternal education††** | | |
| Mean (SD) | 0.8 (±1.3) | 1.2 (±1.5) |
| Missing | 63 (10.5%) | 1 (0.0%) |
| **Diabetes or hypertension in pregnancy** | | |
| Absent | 450 (75.1%) | 1,881 (82.9%) |
| Present | 86 (14.4%) | 387 (17.1%) |
| Missing | 63 (10.5%) | 1 (0.0%) |
| **Gestational age at birth (weeks)** | | |
| Mean (SD) | 38.1 (±3.0) | 38.8 (±2.2) |
| Missing | 9 (1.5%) | 2 (0.1%) |
| **Smoking in pregnancy†††** | | |
| Mean (SD) | 1.0 (±1.6) | 0.6 (±1.2) |
| Missing | 152 (25.4%) | 167 (7.4%) |
| **Treatment for psychiatric disorder** | | |
| Absent | 518 (86.5%) | 2,220 (97.8%) |
| Present | 18 (3.0%) | 48 (2.1%) |
| Missing | 63 (10.5%) | 1 (0.0%) |
| **Maternal alcohol consumption††††** | | |
| Mean (SD) | 5.0 (±1.3) | 4.8 (±1.3) |
| Missing | 63 (10.5%) | 1 (0.0%) |

†Family income: 1=Less than $7,000, 2=$7,000-$11,999, 3=$12,000-$23,999, 4=$24,000-$35,000, 5=$36,000 or more

††Education: 0 = None or `Other`, 1 = Trade certificate or apprenticeship, 2=Professional registration (non-degree), 3=College diploma or degree, 4=University degree

††† 0=None, 1=1 to 5 daily, 2=6 to 10 daily, 3=11 to 15 daily, 4=16 to 20 daily, 5=21 or more per day

††††1=Daily, 2=Several times per week, 3=Approximately once per week, 4, Less than once per week, 5=One binge effort, 6=Never

Supplementary table 2. Additional behavioural assessments used in sensitivity analyses

| | Female (n = 1095) | Male (n = 1174) | P-value |
|---|---|---|---|
| **CBCL Aggression problems age 2** | | | **0.002** |
| Mean (SD) | 17.6 (±10.0) | 19.2 (±10.6) | |
| Missing | 248 (22.6%) | 244 (20.8%) | |
| **TRF Aggression problems age 10** | | | **< 0.0001** |
| Mean (SD) | 2.0 (±4.4) | 5.2 (±7.6) | |
| Missing | 251 (22.9%) | 248 (21.1%) | |
| **TRF Attention problems age 10** | | | **< 0.0001** |
| Mean (SD) | 2.9 (±5.2) | 7.3 (±8.1) | |
| Missing | 251 (22.9%) | 248 (21.1%) | |
| **TRF Social problems age 10** | | | **< 0.0001** |
| Mean (SD) | 1.1 (±2.6) | 1.9 (±3.0) | |
| Missing | 251 (22.9%) | 248 (21.1%) | |

P-value calculated by Willcoxon rank-sum test

Supplementary Table 3. The association between BW and aggressive behaviour in the Raine Study ages 2-17

| | **Model 1** | **Model 2** | **Model 3*** | **Model 4*** |
|---|---|---|---|---|
| **Main effect** | $\boldsymbol{\beta}$: -0.136<br>CI*: [-0.344, 0.0904]<br>SE: 0.091<br>P-value: 0.135 | NA | NA | NA |
| **Baseline effect (females)** | NA | $\boldsymbol{\beta}$: 0.00885<br>CI*: [-0.258, 0.302]<br>SE: 0.117<br>P-value: 0.94 | $\boldsymbol{\beta}$: 0.0118<br>CI*: [-0.404, 0.450]<br>SE: 0.179<br>P-value: 0.948 | $\boldsymbol{\beta}$: -0.0795<br>CI*: [-0.413, 0.264]<br>SE: 0.142<br>P-value: 0.57 |
| **Sex Interaction (males)** | NA | $\boldsymbol{\beta}$: -0.449<br>CI*: [-0.882, -0.0354]<br>SE: 0.177<br>P-value: 0.011 | $\boldsymbol{\beta}$: -0.297<br>CI*: [-0.795, 0.189]<br>SE: 0.21<br>P-value: 0.149 | $\boldsymbol{\beta}$: -0.279<br>CI*: [-0.736, 0.178]<br>SE: 0.191<br>P-value: 0.15 |

* P-value is approximated based on Z-statistic of the bootstrapped SE, but significance is derived from the 98.3 % CI
** Adjusted for age at assessment, maternal BMI, maternal education, maternal psychiatric illness, gestational age at birth, maternal age at birth, maternal smoking during pregnancy, family income during pregnancy, maternal ethnicity and maternal alcohol consumption during pregnancy.
*** Adjusted for age at assessment, maternal BMI, maternal age at birth, maternal smoking during pregnancy, maternal alcohol consumption, family income level and maternal race

Supplementary Table 4. The association between BW and CBCL syndrome scales in the Raine Study ages 5-17 with the exclusion of preterm births using our parsimonious model 4.

| | **Aggressive behaviour**** | **Attention problems***** | **Social problems****** |
|---|---|---|---|
| **Baseline effect (females)** | $\boldsymbol{\beta}$: -0.0893<br>CI*: [-0.471, 0.302]<br>SE: 0.162<br>P-value: 0.58 | $\boldsymbol{\beta}$: 0.00266<br>CI*: [-0.201, 0.209]<br>SE: 0.086<br>P-value: 0.98 | $\boldsymbol{\beta}$: 0.0599<br>CI*: [-0.0748, 0.195]<br>SE: 0.057<br>P-value:0.29 |
| **Sex Interaction (males)** | $\boldsymbol{\beta}$: -0.220<br>CI*: [-0.764, 0.316]<br>SE: 0.226<br>P-value: 0.33 | $\boldsymbol{\beta}$: -0.239<br>CI*: [-0.506, 0.0279]<br>SE: 0.112<br>P-value: 0.032 | $\boldsymbol{\beta}$: -0.175<br>CI*: [-0.334, -0.0093]<br>SE: 0.068<br>P-value: 0.010 |

* P-value is approximated based on Z-statistic of the bootstrapped SE, but significance is derived from the 98.3 % CI
** Adjusted for age at assessment, maternal BMI, maternal age at birth, maternal smoking during pregnancy, maternal alcohol consumption, family income level and maternal race
*** Adjusted for age at assessment, maternal BMI, maternal age at birth, gestational age at birth, maternal smoking during pregnancy, family income level and maternal race
**** Adjusted for age at assessment, maternal BMI, maternal psychiatric illness, maternal age at birth, gestational age at birth, maternal smoking during pregnancy, maternal alcohol consumption during pregnancy and family income level

Supplementary Table 5. The association between BW and CBCL syndrome in the Raine Study age 10 using teacher assessments with the Teacher Report Form in the model 4 regression.

| | **Aggressive behaviour **** | **Attention problems***** | **Social problems ****** |
|---|---|---|---|
| **Baseline effect (females)** | $\boldsymbol{\beta}$: 0.119<br>CI*: [-0.238, 0.482]<br>SE: 0.151<br>P-value: 0.43 | $\boldsymbol{\beta}$: 0.17<br>CI*: [-0.471, 0.803]<br>SE: 0.267<br>P-value: 0.52 | $\boldsymbol{\beta}$: 0.052<br>CI*: [-0.231, 0.334]<br>SE: 0.118<br>P-value: 0.66 |
| **Sex Interaction (males)** | $\boldsymbol{\beta}$: -0.541<br>CI*: [-1.31, 0.218]<br>SE: 0.3199<br>P-value: 0.091 | $\boldsymbol{\beta}$: -0.75<br>CI*: [-1.675, 0.172]<br>SE: 387<br>P-value: 0.052 | $\boldsymbol{\beta}$: -0.343<br>CI*: [-0.720, 0.026]<br>SE: 0.156<br>P-value: 0.0252 |

* P-value is approximated based on Z-statistic of the bootstrapped SE, but significance is derived from the 98.3 % CI
** Adjusted for age at assessment, maternal BMI, maternal age at birth, maternal smoking during pregnancy, family income level and maternal race, maternal alcohol consumption during pregnancy
*** Adjusted for age at assessment, maternal BMI, maternal age at birth, gestational age at birth, maternal smoking during pregnancy, family income level, maternal psychiatric disease and maternal race
**** Adjusted for age at assessment, maternal BMI, maternal age at birth, gestational age at birth, maternal smoking during pregnancy, maternal alcohol consumption during pregnancy, family income level, maternal psychiatric disease and maternal race
